# Predictors of employment attrition in Lebanon during multifaceted crises: The role of chronic diseases – a national cross-sectional study

**DOI:** 10.1101/2025.06.26.25330387

**Authors:** Myriam Dagher, Ali Abboud, Ghada E. Saad, Rita Itani, Hala Ghattas, Stephen J. McCall, WOMENA Study Group

**Affiliations:** Center for Research on Population and Health, Faculty of Health Sciences, American University of Beirut, Beirut, Lebanon; Department of Economics, American University of Beirut, Beirut, Lebanon; Department of Health Promotion, Education, and Behavior, University of South Carolina, Columbia, South Carolina, United States of America

**Author notes:** Department of Health Promotion, Education, and Behavior, University of South Carolina, Columbia, South Carolina, United States of America (Hala Ghattas). Equal contribution (Myriam Dagher, Ali Abboud). Corresponding author: Stephen J. McCall, Center for Research on Population and Health, Faculty of Health Sciences, American University of Beirut, Beirut 1107 2020, Lebanon. The WOMENA Study Group members are listed in the Acknowledgements.

## Abstract

The COVID-19 pandemic and Lebanon’s ongoing economic crisis exacerbated existing inequalities, including workforce disparities. This study identified predictors of employment attrition during Lebanon’s concurrent crises and examined the association between chronic conditions and employment attrition. This cross-sectional study recruited adults aged 19-64 years residing in Lebanon through random digit dialing (5 January – 9 July 2024). Data collected included socio-demographics, household characteristics, employment, and self-reported chronic conditions. The outcome was the loss of paid employment (employment attrition) during the crises. Predictors were identified through LASSO regression and model discrimination and calibration were assessed. Logistic regression models, adjusted for covariates identified through directed acyclic graphs, assessed the association between number and types of chronic conditions and employment attrition. Of 2103 participants employed prior to the onset of the concurrent crises (pre-2020), 72.7% were males, 70.1% were Lebanese, and 14.7% became unemployed during the crises. Predictors of employment attrition were: older age, females, non-Lebanese, married, no formal education, having at least one chronic condition, working in a private or non-governmental organization, and having an oral agreement with employer. The predictive model demonstrated a moderate to good discriminative ability and good calibration. Pre-existing chronic conditions, such as cardiovascular disease (aOR: 2.15; 95% CI, 1.27 to 3.64) and diabetes (aOR: 2.52; 95% CI, 1.43 to 4.45), were independently associated with employment attrition. This study underscores the need to address life-course disparities contributing to job loss and to consider proactive job protections to mitigate workforce disruptions during multiple crises, particularly in contexts where social safety nets are absent.

## Introduction

Decent and inclusive work for all, indicated in the United Nations Sustainable Development Goal 8, is essential for sustainable economic growth and better health outcomes [1]. Despite the reduction in the global unemployment rate to around 5%, inequalities in the labor market still persist globally [2]. These disparities are influenced, and often shaped, by factors rooted in the individual’s life course.

Environmental factors at the macro-level, including the benefits system and economic development, and at the meso-level, such as workplace characteristics and working conditions, affect the capability of working-age individuals to remain in the workforce [3]. For instance, workers in highly physically demanding occupations, such as in construction, experienced shorter working lives by about a year and exited the labor force prematurely [4, 5, 6]. Additionally, poor compensation and temporary contracts have been found to predict employee attrition and turnover using machine learning techniques [7, 8]. Micro-level factors such as sex, age, and education also influence transitions into and out of the labor force. For example, people with primary or secondary education spent more years unemployed than those with tertiary education [9]. In addition, chronic health conditions are an important micro-level barrier to paid employment especially conditions that limit functionality at work, such as cardiovascular disease, diabetes, and musculoskeletal disorders [3, 10, 11].

Lebanon, a low-middle-income country (LMIC) in the Middle East and North Africa (MENA) region, is characterized by low labor force participation, especially among women [1]. From late 2019 to 2023, Lebanon has been grappling with concurrent crises; from a protracted political and economic crises to the COVID-19 pandemic and the Beirut Port explosion. The labor underutilization rate in the country dramatically increased from around 16% between 2018 and 2019 to 50% in 2022, accompanied by a gender gap in employment [1]. The crises have added another layer of complexity to the already existing workforce disparities and health challenges in the country, with expectations of further decreasing employment in the coming years [12]. Studies exploring factors associated with unemployment in LMIC settings, particularly in the context of overlapping crises, are scarce. One recent study from five LMICs of the MENA region – Egypt, Jordan, Morocco, Sudan, and Tunisia – explored changes in employment associated with care responsibilities during the pandemic, focusing solely on women [13]. Thus, this study aimed to explore the predictors of employment attrition during Lebanon’s concurrent crises and examined the association between pre-existing chronic conditions and employment attrition.

## Methods

### Study design and setting

This was a national cross-sectional study conducted as part of a larger research project entitled “Identifying opportunities to improve the lived experience and health of working women in the MENA: from COVID to recovery-WOMENA.” The main aim of the project was to examine trends in the gender distribution of labor force participation in the MENA region and how these trends impacted health and well-being, with a particular focus on the impact of the COVID-19 pandemic. This study represented a component of the larger research project and specifically aimed to understand the intersection between labor force participation and health in Lebanon. The study protocol was reviewed and approved by the [name of university] Social and Behavioral Sciences Institutional Review Board [Reference: SBS-2023-0182] and implemented in compliance with its ethical guidelines. As the data collection was conducted through a telephone survey, oral consent was obtained verbally from all participants prior to their involvement in the study and the necessary measures have been taken to protect participants’ privacy.

### Sampling and study population

Participants were recruited through Random Digit Dialing (RDD) between January 2024 and July 2024. Random numbers were generated to construct potential phone numbers using the 11-digit structure for mobile phone numbers in Lebanon. These numbers had the first three digits corresponding to the international country calling code for Lebanon (961), followed by two digits indicating the country’s mobile network operators (03, 70, 71, 76, 78, 79, 81), and the remaining six numbers were randomly generated. Each number generated was dialed with a maximum of two call attempts. If respondents were unable to answer or complete the survey at the time of the initial call, follow-up call appointments were scheduled for a more convenient time.

Data were collected by trained data collectors in the participants’ mother tongues and data entry was performed using SurveyCTO software [14]. A comprehensive monitoring plan and standardized data quality procedures were implemented alongside data collection, with weekly monitoring to identify and address systematic errors in real-time. As part of the process, 5% of surveys were recorded and cross-checked against the actual data entered, resulting in an average error rate of 0.59%, which was corrected through recording replays or follow-up calls, when necessary.

Respondents completed a set of eligibility screening questions. The survey was restricted to working age adults (age 19-64 years), who were permanently residing in Lebanon at the time of the survey. Respondents who were on a visit or temporary stay were not eligible to participate in the survey. There were no restrictions based on citizenship or legality of residency status. Employed women were oversampled given the low female labor force participation in the population.

### Measures

#### Outcome measures

The outcome of interest in this study was the loss of paid employment (employment attrition) during the peak period of the concurrent pandemic and economic crisis in Lebanon (2020–2023) [15, 16]. The employment module included current and retrospective employment questions. Participants were asked about the start and end date of every job they had in the past 10 years. The data allowed the construction of the outcome variable, employment attrition; a binary variable assigned the value of 1 if the individual “left employment”, and 0 if the individual “maintained or remained employed” during the crises (between 2020 and 2023). The sample was therefore naturally restricted to those who were employed prior to the onset of the crises or before 2020 (n=2103). Additionally, participants who left employment were asked about the reasons for their work discontinuation.

#### Candidate predictors and data

Ten candidate predictors for employment attrition were identified through the literature and included in the model development. These predictors were age, sex, nationality, marital status, education, urbanization of living environment, number of children, and number of pre-existing chronic conditions, including cardiovascular disease (CVD), diabetes, and musculoskeletal disorders. These chronic conditions were selected as they have been shown to be associated with the risk of early retirement or increase the risk of reduced labor market participation [17, 18, 19]. The candidate predictors also included job sector and contractual agreement prior to the crises. The degree of urbanization of living environment was assessed through a single-item self-report measure “Please indicate how urban your living environment is on a 7-point scale from (1) not urban at all to (7) very urban.” The responses were then presented as binary, with a score of 6 distinguishing *extremely urbanized* from *not urbanized* or *strongly urbanized areas* [20]. Self-reported chronic conditions were measured by asking participants if they had ever been diagnosed or informed by a healthcare professional about any of the following chronic conditions or diseases: CVD, diabetes, musculoskeletal disorders, hypertension, chronic respiratory disease, chronic kidney disease, and neurodegenerative conditions. The onset of diagnosis recalled by participants, enabled the distinction between pre-crises (referred to as pre-existing) and current chronic conditions, as the study’s secondary aim is to understand how pre-existing chronic conditions might have influenced employment attrition during the crises.

The variable with the highest proportion of missing values accounted for 1.99% of the dataset. Missing data were missing completely at random using the Little’s test and a complete case analysis was performed [21].

### Statistical analysis

Absolute frequencies and weighted proportions were presented for categorical variables, and medians with their interquartile range (IQR) for continuous variables. Weighted odds ratios (ORs) alongside their 95% confidence intervals (CIs) were calculated using unadjusted logistic regression models to examine the odds of employment attrition for each independent variable. All candidate predictors were categorical except age and number of children, which had a linear association with employment attrition. Variables with P-values less than 0.05 were considered statistically significant. Sampling and post-calibration weights were calculated and applied to the analysis to allow for national estimates.

Least Absolute Shrinkage and Selection Operator (LASSO) logistic regression model was used to identify the predictors of employment attrition during the concurrent crises for the study sample. All candidate predictors were entered for model development, which was internally validated using 10-fold cross-validation. Separate logistic regressions were run on selected predictors to produce ORs and 95% CIs.

The final model performance was assessed through its discrimination and calibration abilities [22]. C-Statistic/Area Under the Receiver Operating Curve (AUC), ranges from 0.5 to 1 with an index of 1 indicating perfect discrimination ability or how well predictions differentiated participants who experienced employment attrition from those who did not. The evaluation of the model calibration (agreement between predicted and observed risk) was assessed through a calibration plot and calibration slope (C-Slope), and calibration-in-the-large (CITL). A good model calibration in future datasets is illustrated by a diagonal line with an intercept of 0 and a slope of 1 [23]. All analyses were conducted using Stata/SE statistical software version 18 (StataCorp LLC).

### Association between pre-existing chronic conditions and employment attrition

A secondary analysis explored the association between the number and types of chronic conditions (exposures) and employment attrition during the crises (outcome). The outcome employment attrition was analyzed as a binary variable. The exposures included having, prior to the crises: i) at least one chronic condition from all listed chronic conditions, and ii) at least one of the chronic conditions that have been shown to be associated with the risk of early retirement or increase the risk of reduced labor market participation (CVD, diabetes, musculoskeletal disorders). The former exposures were treated as binary; coded as 0 for no pre-existing chronic condition(s) and 1 for having at least one pre-existing chronic condition. Similarly, each of the following conditions were treated individually as binary exposures; those with a pre-existing history of CVD, diabetes, musculoskeletal disorders, hypertension, chronic respiratory disease, chronic kidney disease, and neurodegenerative conditions.

To account for confounders, a Directed Acyclic Graph (DAG) was constructed using DAGitty software (dagitty.net) [24, 25]. The DAG represented a conceptual model that informed our understanding of potential pathways through which having chronic conditions could lead to an increased risk of exiting paid employment during the concurrent crises in Lebanon. It included key variables shaping this relationship and their interconnections, as informed by the literature in the context of the study area [26, 27, 28, 29, 30]. The initial DAG was validated by three members of the study team and underwent minor modifications based on consensus (S2 Figure).

The minimal sufficient adjustment set included age, sex, marital status, job sector before the crises, and job income before the crises (USD per year, used as a continuous variable). For each exposure, separate logistic regression models were used to assess the association between the number or type of chronic conditions and employment attrition after adjusting for the suggested covariates. Weighted unadjusted and adjusted ORs were calculated with their associated 95% CIs.

## Results

A total of 97608 phone numbers were contacted through RDD, of whom 7372 responded and were eligible. Among them, 4725 completed the full survey. After excluding 1432 participants who started employment in 2020 or later, 693 who never worked, 195 whose last job ended in and before 2014, and 101 who were uncertain or unwilling to disclose their job start date; a total of 2304 participants were employed before the crises (2020–2023). The final study sample for analysis comprised 2103 participants who either remained employed after 2023 or left employment during the crises (Fig 1).

**Fig 1.**
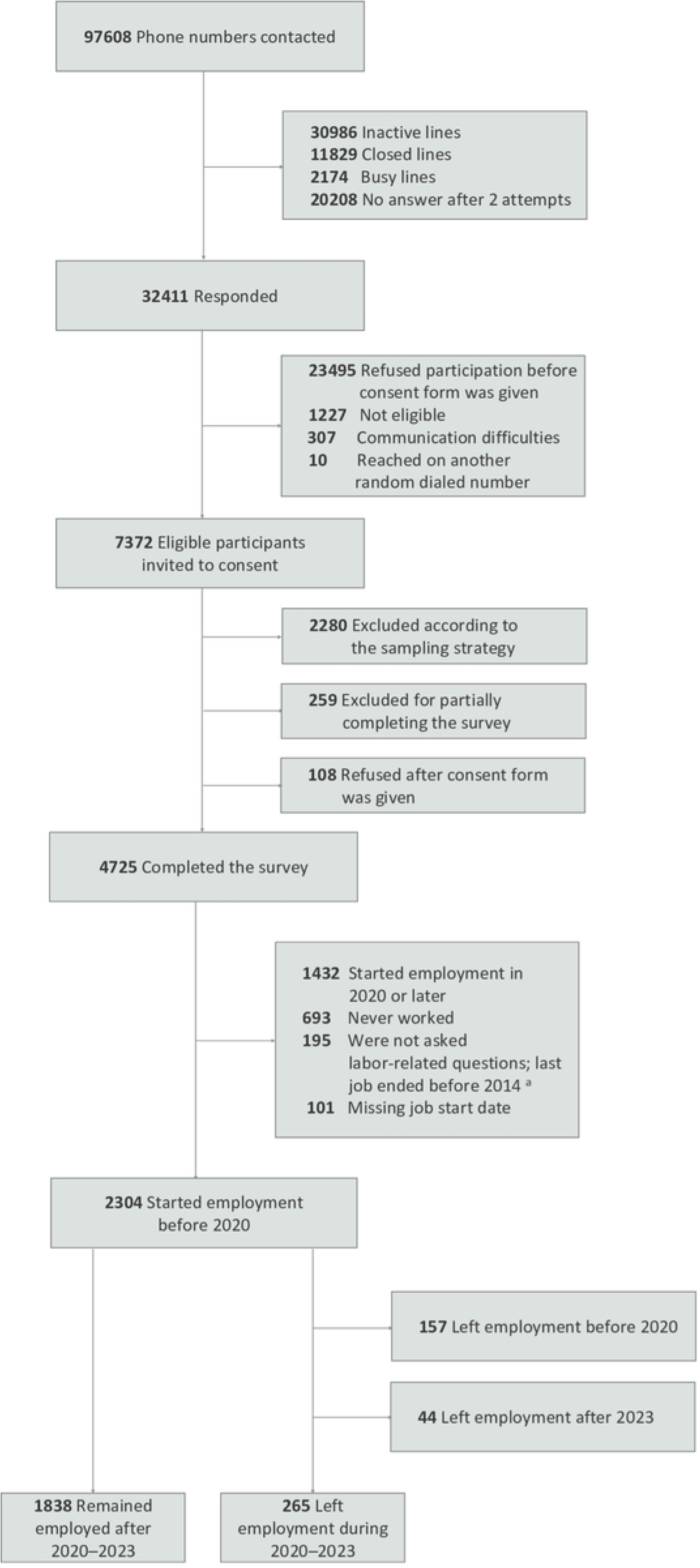
Flow Diagram of Contacted Phone Numbers and Study Sample (n = 2103). ^a^ When collecting retrospective data on employment, we did not ask about employment characteristics and history for individuals whose last job falls beyond 10-year recall period (i.e., whose last job ended before 2014).

The median age of the participants in the study sample was 40 years (IQR: 33-49 years); 1466 participants (72.7%) were males and 1311 (70.1%) were Lebanese. Of the study sample, 265 (14.7%) participants experienced employment attrition during the crises, while 1838 (85.3%) remained employed after the crises (Table 1). The reasons reported for leaving work during the crises included illness, disability, or injury (23.0%); ending economic activity through resignation or retirement (18.1%); temporary or seasonal job (17.7%); inappropriate working conditions related to working hours, salary, work environment, or logistical challenges accessing the workplace (17.1%); family sickness or responsibilities such as caregiving, marriage, or pregnancy (16.8%); and dismissal or redundancy (11.1%) (S1 Figure). Among the study sample, 310 (15.4%) participants reported having at least one pre-existing chronic condition, including 7.9% who had musculoskeletal disorders, 6.6% CVD, and 5.1% diabetes (Table 1).

**Table 1.** Characteristics of the Study Sample Stratified by Employment Attrition During the Concurrent Crises in Lebanon (2020-2023) (n = 2103).

|  | Participants, No. (Weighted %)<sup>a</sup> |  |  | Weighted<br>Unadjusted Odds<br>Ratio (95% CI)<sup>b</sup> | *P* values |
| --- | --- | --- | --- | --- | --- |
|  | Total<br>(n = 2103) | Remained employed<br>after the crises<br>(n = 1838) | Left employment<br>during the crises<br>(n = 265) |  |  |
| Age, median (IQR), years | 40 (33-49) | 40 (33-49) | 40 (32-51) | 1.00 (0.99 to 1.16) | .676 |
| Sex |  |  |  |  |  |
| Men | 1466 (72.7) | 1293 (88.9) | 173 (11.1) | 1 [Reference] |  |
| Women | 637 (27.3) | 545 (75.5) | 92 (24.5) | 2.61 (1.98 to 3.44) | <.001 |
| Nationality |  |  |  |  |  |
| Lebanese | 1311 (70.1) | 1178 (87.1) | 133 (12.9) | 1 [nce] |  |
| Non-Lebanese | 792 (29.9) | 660 (81.0) | 132 (19.0) | 1.58 (1.21 to 2.05) | .001 |
| Marital status |  |  |  |  |  |
| Married | 1588 (75.2) | 1379 (84.9) | 209 (15.1) | 1 [Reference] |  |
| Non-married <sup>c</sup> | 515 (24.8) | 459 (86.3) | 56 (13.7) | 0.89 (0.65 to 1.24) | .499 |
| Education |  |  |  |  |  |
| Never attended school | 67 (2.9) | 48 (65.4) | 19 (34.6) | 1 [Reference] |  |
| Formal or technical education | 1500 (70.9) | 1296 (84.6) | 204 (15.4) | 0.34 (0.19 to 0.60) | <.001 |
| Higher education | 536 (26.2) | 494 (89.1) | 42 (10.9) | 0.23 (0.12 to 0.44) | <.001 |
| Urbanization of living environment |  |  |  |  |  |
| Not urbanized/strongly urbanized | 1642 (80.0) | 1420 (84.5) | 222 (15.5) | 1 [Reference] |  |
| Extremely urbanized | 439 (20.0) | 398 (88.1) | 41 (11.9) | 0.74 (0.51 to 1.06) | .102 |
| Missing | 22 | 20 | 2 | NA | NA |
| Children, median (IQR), No. | 2 (1-4) | 2 (1-4) | 2 (1-4) | 1.04 (0.98 to 1.10) | .249 |
| Pre-existing <sup>d</sup> chronic conditions<br>linked with unemployment, No.<sup>e</sup> |  |  |  |  |  |
| 0 | 1786 (84.6) | 1571 (85.9) | 215 (14.1) | 1 [Reference] |  |
| ≥ 1 | 310 (15.4) | 260 (81.2) | 50 (18.8) | 1.41 (1.00 to 2.00) | .053 |
| Missing | 7 | 7 | 0 | NA | NA |
| Cardiovascular disease, pre-existing |  |  |  |  |  |
| No | 1961 (93.4) | 1723 (85.8) | 238 (14.2) | 1 [Reference] |  |
| Yes | 125 (6.6) | 98 (75.6) | 27 (24.4) | 1.95 (1.23 to 3.09) | .004 |
| Missing | 17 | 17 | 0 |  |  |
| Diabetes, pre-existing |  |  |  |  |  |
| No | 1995 (94.9) | 1752 (85.7) | 243 (14.3) | 1 [Reference] |  |
| Yes | 94 (5.1) | 73 (75.7) | 21 (24.3) | 1.93 (1.15 to 3.24) | .013 |
| Missing | 14 | 13 | 1 | NA | NA |
| Musculoskeletal disorders, pre-existing |  |  |  |  |  |
| No | 1924 (92.1) | 1684 (85.4) | 240 (14.6) | 1 [Reference] |  |
| Yes | 167 (7.9) | 142 (82.2) | 25 (17.8) | 1.27 (0.79 to 2.03) | .319 |
| Missing | 12 | 12 | 0 | NA | NA |
| Job sector, pre-2020 <sup>d</sup> |  |  |  |  |  |
| Government | 271 (14.1) | 254 (92.0) | 17 (8.0) | 1 [Reference] |  |
| Private business | 978 (46.6) | 829 (82.5) | 149 (17.5) | 2.45 (1.44 to 4.17) | .001 |
| Private household | 60 (2.4) | 56 (91.0) | 4 (9.0) | 1.14 (0.35 to 3.71) | .830 |
| Non-governmental institution | 75 (3.6) | 61 (75.1) | 14 (24.9) | 3.81 (1.75 to 8.31) | .001 |
| Freelance | 715 (33.3) | 635 (87.0) | 80 (13.0) | 1.72 (0.99 to 3.00) | .056 |
| Missing | 4 | 3 | 1 | NA | NA |
| Contractual agreement, pre-2020 <sup>d</sup> |  |  |  |  |  |
| Written | 489 (25.1) | 449 (89.4) | 40 (10.6) | 1 [Reference] |  |
| Oral | 921 (42.4) | 756 (79.3) | 165 (20.7) | 2.20 (1.51 to 3.20) | <.001 |
| Independent contractor <sup>f</sup> | 651 (32.5) | 595 (89.6) | 56 (10.4) | 0.97 (0.63 to 1.50) | .897 |
| Missing | 42 | 38 | 4 | NA | NA |

<sup>a</sup> Column percentages are given for the overall sample (n = 2103). Row percentages are used to show the distribution of the variables stratified by the outcome. Percentages are presented in parentheses. 95% Confidence Intervals (CIs) are shown in parentheses with “to” separating the range. The reference category is indicated using square brackets (e.g., 1 [Reference]).
<sup>b</sup> Dependent (outcome) variable is individuals who became unemployed any time during the concurrent crises (2020-2023) compared to those who were still employed after the crises.
<sup>c</sup> Non-married includes participants who are single, engaged, divorced/separated, or widowed.
<sup>d</sup> Pre-existing or pre-2020 refers to conditions or circumstances that were present before the onset of the concurrent crises, prior to the year 2020.
<sup>e</sup> Cardiovascular disease, diabetes, and musculoskeletal disorders have been shown to be associated with the risk of early retirement or increase the risk of reduced labor market participation.
<sup>f</sup> Independent contractor includes owners, partners, and own account workers.

The weighted unadjusted ORs and 95% CIs of the association between employment attrition during the crises and each characteristic are reported in Table 1. Among the study sample, women were significantly more likely than men to leave employment during the crises (OR, 2.61; 95% CI, 1.98 to 3.44), as were non-Lebanese (OR, 1.58; 95% CI, 1.21 to 2.05), those who had pre-existing CVD, diabetes, or musculoskeletal disorders (OR, 1.41; 95% CI, 1.00 to 2.00), those who worked in private businesses (OR, 2.45; 95% CI, 1.44 to 4.17) or in non-governmental institutions (OR, 3.81; 95% CI, 1.75 to 8.31) prior to the crises, and those who had an oral agreement with their employer prior to the crises (OR, 2.20; 95% CI, 1.51 to 3.20).

Additionally, participants living in extremely urbanized areas had lower odds of employment attrition compared to those living in the least urbanized areas (OR, 0.74; 95% CI, 0.51 to 1.06). Those with formal or technical education (OR, 0.34; 95% CI, 0.19 to 0.60) or higher education (OR, 0.23; 95% CI, 0.12 to 0.44) were less likely than those who never attended school to become unemployed during the crises.

### Predictors of employment attrition and model performance

Table 2 presents the penalized coefficients, adjusted ORs, and their corresponding 95% CIs. Ten predictors of employment attrition were identified including age, sex, nationality, marital status, education, urbanization of living environment, number of children, number of chronic conditions (CVD, diabetes, or musculoskeletal disorders), job sector prior to the crises, and contractual agreement prior to the crises. The predictors’ coefficients indicated that older age, females, having non-Lebanese nationality, being married, never having attended school, having at least one pre-existing CVD, diabetes, or musculoskeletal conditions, working in a private business or non-governmental institution, and having an oral agreement with employer predicted employment attrition during the concurrent crises in Lebanon.

**Table 2.** Predictors of Employment Attrition in the Study Sample and Adjusted Odds Ratios from Logistic Regression (n = 2027).

| Predictor variables included in the final model | Codes | Penalized Coefficients <sup>a</sup> | Adjusted Odds Ratio (aOR) (95% CI) <sup>b</sup> |
| --- | --- | --- | --- |
| Age (years) |  | 0.14 | 1.01 (1.00 to 1.03) |
| Sex |  |  |  |
| Men | 0 |  | 1 [Reference] |
| Women | 1 | 0.57 | 1.79 (1.32 to 2.43) |
| Nationality |  |  |  |
| Lebanese | 0 |  | 1 [Reference] |
| Non-Lebanese | 1 | 0.37 | 1.47 (1.06 to 2.05) |
| Marital status |  |  |  |
| Married | 0 | 0.21 | 1 [Reference] |
| Non-married <sup>c</sup> | 1 |  | 0.80 (0.55 to 1.16) |
| Education |  |  |  |
| Never attended school | 0 | 0.77 | 1 [Reference] |
| Formal or technical education | 1 | - | 0.46 (0.25 to 0.83) |
| Higher education | 2 | -0.54 | 0.26 (0.13 to 0.53) |
| Urbanization of living environment |  |  |  |
| Not urbanized/strongly urbanized | 0 |  | 1 [Reference] |
| Extremely urbanized | 1 | -0.58 | 0.55 (0.38 to 0.80) |
| Children number, No. |  | -0.04 | 0.95 (0.88 to 1.03) |
| Pre-existing <sup>d</sup> chronic conditions linked with unemployment, No. <sup>e</sup> |  |  |  |
| 0 | 0 |  | 1 [Reference] |
| ≥ 1 | 1 | 0.39 | 1.48 (1.02 to 2.15) |
| Job sector, pre-2020 <sup>d</sup> |  |  |  |
| Government | 0 | -0.29 | 1 [Reference] |
| Private business | 1 | 0.21 | 1.68 (0.93 to 2.99) |
| Private household | 2 | -0.96 | 0.50 (0.15 to 1.98) |
| Non-governmental institution | 3 | 0.48 | 2.20 (0.90 to 4.70) |
| Freelance | 4 | - | 1.35 (0.70 to 2.63) |
| Contractual agreement, pre-2020<sup>d</sup> |  |  |  |
| Written | 0 | - | 1 [Reference] |
| Oral | 1 | 0.66 | 1.93 (1.23 to 3.04) |
| Independent contractor<sup>f</sup> | 2 | -0.11 | 0.88 (0.51 to 1.52) |
| Intercept |  | -3.16 | 0.08 (0.03 to 0.23) |

<sup>a</sup> Penalized coefficient obtained after internal validation using 10-fold cross-validation.
<sup>b</sup> Dependent (outcome) variable is individuals who became unemployed any time during the concurrent crises (2020-2023) compared to those who were still employed after the crises.
<sup>c</sup> Non-married includes participants who are single, engaged, divorced/separated, or widowed.
<sup>d</sup> Pre-existing or pre-2020 refers to conditions or circumstances that were present before the onset of the concurrent crises, prior to the year 2020.
<sup>e</sup> Cardiovascular disease, diabetes, and musculoskeletal disorders have been shown to be associated with the risk of early retirement or increase the risk of reduced labor market participation.
<sup>f</sup> Independent contractor includes owners, partners, and own account workers.

The model demonstrated a moderate to good discriminative ability with an AUC of 0.68 (95% CI, 0.65 to 0.72). The model also showed good calibration with a C-Slope of 1.00 (95% CI, 0.80 to 1.20) (Fig 2).

**Fig 2.**
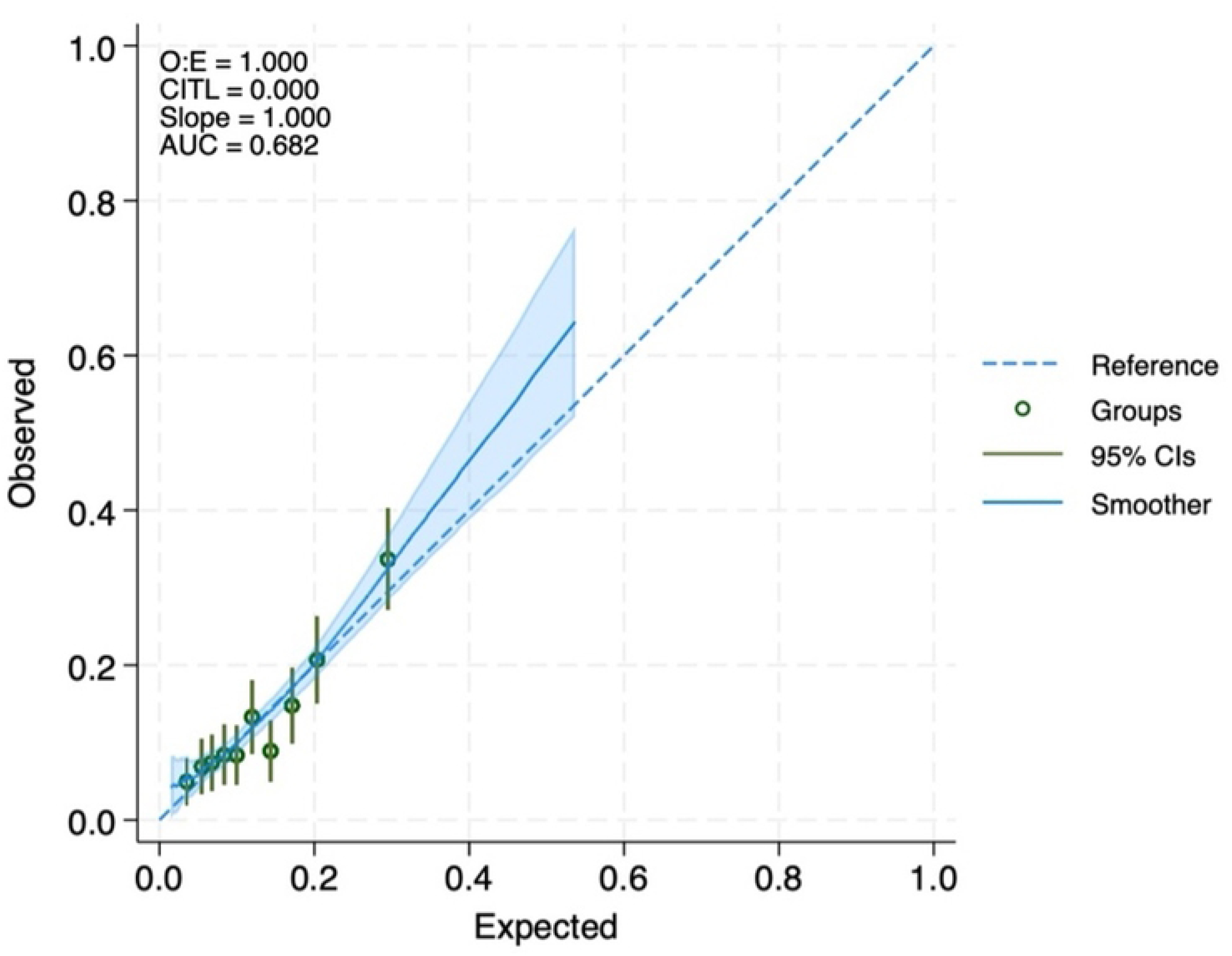
Calibration Plot of the Prediction Model for Employment Attrition During the Concurrent Crises in Lebanon. The x-axis represents the expected probabilities estimated by the model and the y-axis represents the observed probabilities. The dashed diagonal line represents perfect calibration. Whiskers, 95% confidence intervals (CIs); O.E, observed-to-expected ratio; CITL, calibration-in-the-large; and AUC, the Area Under the Receiver Operating Curve.

### Chronic conditions and employment attrition

The weighted unadjusted and adjusted ORs for employment attrition by pre-existing chronic condition status are presented in Table 3. Participants with at least one of the following pre-existing chronic conditions – CVD, diabetes, or musculoskeletal disorders – had significantly higher odds of losing employment during the crises in Lebanon (aOR: 1.57; 95% CI, 1.05 to 2.34), compared to those without any of these pre-existing conditions. When considering each of these chronic conditions separately, participants with pre-existing CVD (aOR: 2.15; 95% CI, 1.27 to 3.64) or diabetes (aOR: 2.52; 95% CI, 1.43 to 4.45) had higher odds of employment attrition compared to participants without these conditions.

**Table 3.**
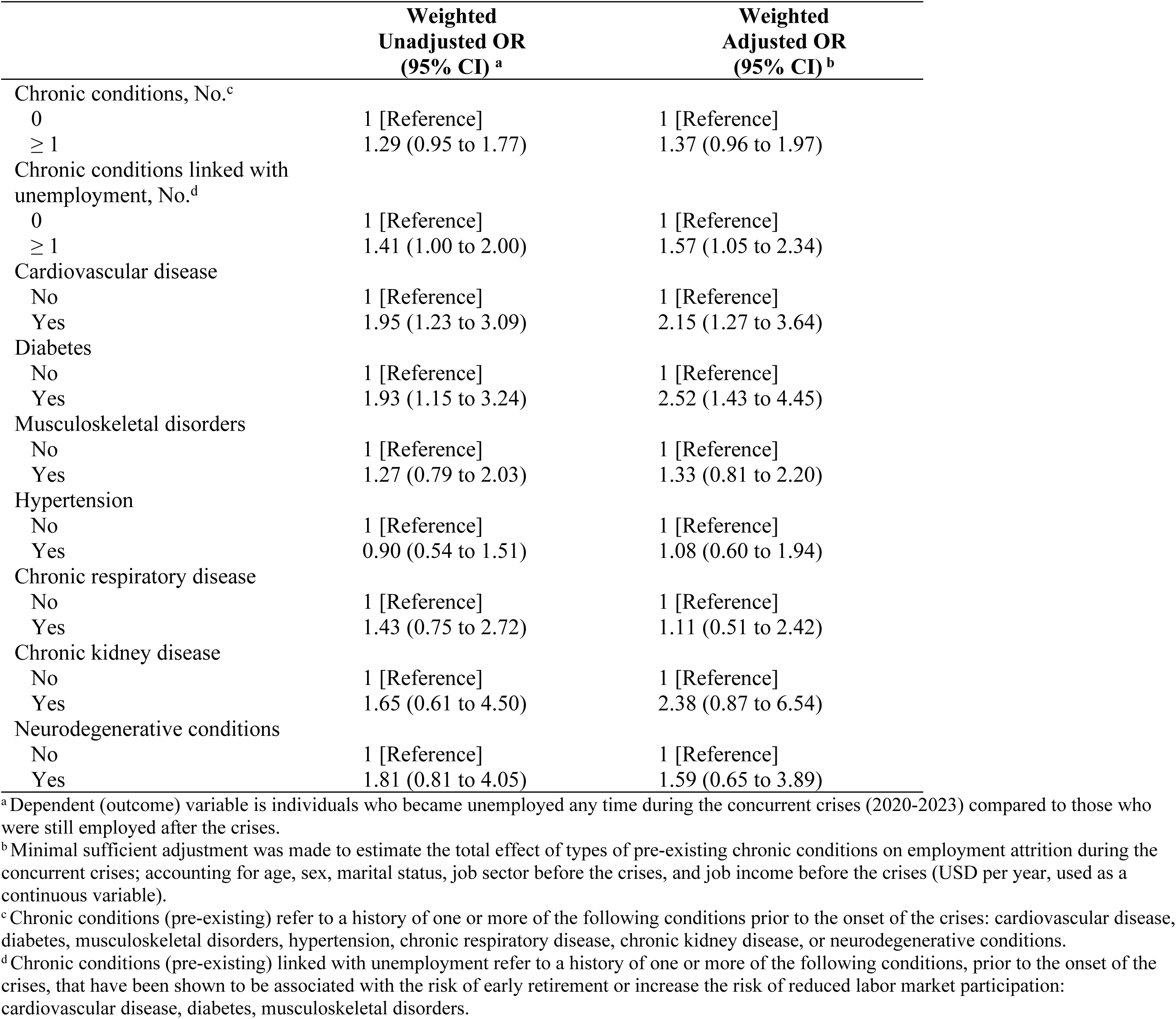
Logistic Regression Models for the Association Between Employment Attrition during the concurrent crises in Lebanon (2020-2023) and Pre-existing Chronic Conditions.

|  | Weighted<br>Unadjusted OR<br>(95% CI) <sup>a</sup> | Weighted<br>Adjusted OR<br>(95% CI) <sup>b</sup> |
| --- | --- | --- |
| Chronic conditions, No.<sup>c</sup> |  |  |
| 0 | 1 [Reference] | 1 [Reference] |
| ≥ 1 | 1.29 (0.95 to 1.77) | 1.37 (0.96 to 1.97) |
| Chronic conditions linked with<br>unemployment, No.<sup>d</sup> |  |  |
| 0 | 1 [Reference] | 1 [Reference] |
| ≥ 1 | 1.41 (1.00 to 2.00) | 1.57 (1.05 to 2.34) |
| Cardiovascular disease |  |  |
| No | 1 [Reference] | 1 [Reference] |
| Yes | 1.95 (1.23 to 3.09) | 2.15 (1.27 to 3.64) |
| Diabetes |  |  |
| No | 1 [Reference] | 1 [Reference] |
| Yes | 1.93 (1.15 to 3.24) | 2.52 (1.43 to 4.45) |
| Musculoskeletal disorders |  |  |
| No | 1 [Reference] | 1 [Reference] |
| Yes | 1.27 (0.79 to 2.03) | 1.33 (0.81 to 2.20) |
| Hypertension |  |  |
| No | 1 [Reference] | 1 [Reference] |
| Yes | 0.90 (0.54 to 1.51) | 1.08 (0.60 to 1.94) |
| Chronic respiratory disease |  |  |
| No | 1 [Reference] | 1 [Reference] |
| Yes | 1.43 (0.75 to 2.72) | 1.11 (0.51 to 2.42) |
| Chronic kidney disease |  |  |
| No | 1 [Reference] | 1 [Reference] |
| Yes | 1.65 (0.61 to 4.50) | 2.38 (0.87 to 6.54) |
| Neurodegenerative conditions |  |  |
| No | 1 [Reference] | 1 [Reference] |
| Yes | 1.81 (0.81 to 4.05) | 1.59 (0.65 to 3.89) |

<sup>a</sup> Dependent (outcome) variable is individuals who became unemployed any time during the concurrent crises (2020-2023) compared to those who were still employed after the crises.
<sup>b</sup> Minimal sufficient adjustment was made to estimate the total effect of types of pre-existing chronic conditions on employment attrition during the concurrent crises; accounting for age, sex, marital status, job sector before the crises, and job income before the crises (USD per year, used as a continuous variable).
<sup>c</sup> Chronic conditions (pre-existing) refer to a history of one or more of the following conditions prior to the onset of the crises: cardiovascular disease, diabetes, musculoskeletal disorders, hypertension, chronic respiratory disease, chronic kidney disease, or neurodegenerative conditions.
<sup>d</sup> Chronic conditions (pre-existing) linked with unemployment refer to a history of one or more of the following conditions, prior to the onset of the crises, that have been shown to be associated with the risk of early retirement or increase the risk of reduced labor market participation: cardiovascular disease, diabetes, musculoskeletal disorders.

Participants with at least one of the broader range of chronic conditions showed increased odds of employment attrition compared to those without any of these conditions. Similarly, when examining each of these conditions individually; those with musculoskeletal disorders, hypertension, chronic respiratory disease, chronic kidney disease, or neurodegenerative conditions were associated with higher odds of employment attrition. However, these associations were not statistically significant.

## Discussion

This study explored predictors of employment attrition among 19 to 64 years old individuals residing in Lebanon during the concurrent pandemic and economic crises, between 2020 and 2023. Being of older age, females, non-Lebanese, married, having no formal education, having at least one chronic condition, working in a private business or non-governmental institution, and having an oral agreement with employer were predictors of employment attrition during times of crises in Lebanon. The predictive model demonstrated a moderate to good discriminative ability and good calibration. In addition, pre-existing chronic conditions, such as CVD and diabetes, were also independently associated with employment attrition.

Micro-level predictors identified in this study align with a systematic review of global evidence, which found age, gender, ethnicity, and education as potential moderators of the COVID-19 pandemic impact on individual labor market outcomes [26] and correlates of labor force instability [31]. Additionally, studies examining predictors of employment outcomes have consistently shown that older age, female sex, and lower educational attainment increased the risk of unemployment [10, 32]. This trend suggests that, when these structural vulnerabilities are already embedded at the population level, the risk of early exit from paid employment may be further exacerbated by the added complexity of crises and economic instability. In some countries of the MENA region such as in Sudan, married women, particularly those with children, were more likely to leave employment during the pandemic [13]. Our results were consistent with this trend, which could be explained by the increased care burden women had to bear as a result of the COVID-19 pandemic lockdown measures, creating additional caregiving responsibilities for married women.

In addition, this study shows meso-level factors, such as working in a private business or non-governmental institution, as predictors of employment attrition, aligning with previous research that demonstrated the larger impact of the pandemic and economic distress on private sector workers compared to workers in other sectors, globally and in most of the MENA countries [13, 26]. These sectors could have been hit harder than others due to the reduced consumer or service demands during times of crises and increased sensitivity of private-sector workers layoffs during economic downturns compared to the public sector [13, 26].

Furthermore, the assessment of the association between employment attrition and the number and type of chronic conditions showed that having pre-existing CVD or diabetes was significantly associated with higher odds of job loss during the crises in Lebanon. Our findings are consistent with previous research that attributed CVD and diabetes mellitus type 2 to lower labor force participation and lower likelihood of receiving disability benefits [3, 10, 11]. Health conditions, especially if poorly managed, can affect employment trajectories due to disease prognosis interfering with the ability to work and have a good quality of life [33].

While there was an increased likelihood of employment attrition when exploring each type of chronic condition, the association did not reach statistical significance for the remaining chronic conditions – musculoskeletal disorders, hypertension, chronic respiratory disease, chronic kidney disease, and neurodegenerative conditions. The literature is inconsistent with regards to these conditions; for example, some studies showed that neurological diseases and chronic obstructive pulmonary disease predicted loss of employment or increased the likelihood of receiving disability benefits [10, 34], while other studies did not find significant associations [35, 36]. This discrepancy could be explained by small sample sizes that are insufficient to detect this association.

Furthermore, the relationship between chronic conditions and employment is intricate. Employment can secure access to healthcare through employer-sponsored insurance and disposable income for out-of-pocket spending, making employment more desirable for people with chronic conditions (Pinto et al., 2018). Conversely, having a chronic condition and its accompanying physical or mental hardship can make participation in the labor market more challenging, especially during crises, further complicating this relationship by increasing the likelihood of maintaining a job. This discrepancy may also be attributed to differences in disease prognosis and how it interferes with work ability, which depend on types of jobs involved and context-specific management strategies. For instance, in some countries such as in Germany, employees with disabilities are protected by national law against dismissal and provided with structural support [37].

This study recognized some limitations. The predictive model had a moderate to good discriminative ability, which may be attributed to missing predictors, yet it remains comparable to prediction models of involuntary exit from paid employment applied in other contexts (C-indexes ranging from 0.70 to 0.85) [10]. Despite using the DAG to delineate temporal order between pre-existing chronic conditions and employment attrition, the cross-sectional nature of the study limits the strength of conclusions that can be made. Also, the outcome of this study was self-reported, nonetheless, the study classified individuals based on their job history dates to reduce information bias. In addition, rigorous quality checks were conducted, and 5% of the surveys were recorded with an average error rate of 0.59% only.

This nationally representative study fills an important gap in a context marked by multiple crises, identifying individuals who experienced heightened vulnerability to exiting the workforce. The findings underscore the importance of addressing disparities embedded within the life course of individuals that contribute to job attrition and emphasize the necessity for proactive job protections to mitigate workforce disruptions during times of crises, particularly in LMIC contexts where social safety nets are absent or limited in capacity. While this cross-sectional study provides valuable insights, longitudinal studies in LMICs are necessary to investigate this relationship further.

## Data Availability

The data used in this study are not publicly available to comply with privacy and ethical considerations. The data is available upon request from the Center for Research on Population and Health at the American University of Beirut.

## Glossary

AUC: Area Under the Receiver Operating Curve
C-index: Concordance index
C-slope: Calibration Slope
CI: Confidence intervals
CITL: Calibration-in-the-large
CVD: Cardiovascular disease
DAG: Directed Acyclic Graph
ILO: International Labour Organization
IQR: Interquartile range
LASSO: Least Absolute Shrinkage and Selection Operator
LMIC: Low-middle-income country
MENA: Middle East and North Africa
aOR: Adjusted Odds ratio
OR: Odds ratio
UNDP: United Nations Development Programme
RDD: Random Digit Dialing

## Acknowledgements

We would like to acknowledge the International Development Research Centre (IDRC) – Canada for funding the research making its implementation possible. We also acknowledge the assistance of B.O.T (Bridge. Outsource. Transform) in data collection. We would like to thank the members of the WOMENA Study Group for their time and contribution to this study: Myriam Dagher, Ali Abboud, Ghada Saad, Rita Itani, Jocelyn DeJong, Malak Ghezzawi, Nisreen Salti, Sasha Fahme, and Serena Canaan, Hala Ghattas, Stephen McCall.

## Supporting Information

**S1 Figure. Primary Reasons for Leaving Work During the Concurrent Crises Among Participants Who Experienced Employment Attrition During the Crises (n = 265).** Dismissed or made redundant refers to participants who were involuntarily terminated by their employer due to either (i) dismissal, involving conduct or capability issues such as poor performance, misconduct, or violation of company policies; or (ii) redundancy, due to company-related factors such as restructuring, downsizing, technological advancements, or decline in demands. Ending economic activity refers to participants who resigned or retired. Inappropriate working conditions includes working hours, salary, work environment, or logistical challenges accessing the workplace. Family sickness/responsibilities include participants who left their work due to family illness, caregiving, marriage, or pregnancy.

**S2 Figure. Directed Acyclic Graph (DAG) of the Proposed Causal Model Between the Number and Types of Chronic Conditions and Employment Attrition During the Concurrent Crises in Lebanon.** DAG was created using http://www.dagitty.net/. SES, socioeconomic status; Pre-2020, before the year 2020.

